# EFFECTS OF HAZARDOUS ALCOHOL AND DRUG USE ON ANTIRETROVIRAL ADHERENCE AND HIV VIRAL SUPPRESSION: A MEDIATION ANALYSIS

**DOI:** 10.1101/2021.10.19.21265220

**Authors:** Tesfaye S. Moges, Edward R. Cachay, Huifang Qin, Laura Bamford, David J. Grelotti, Wm. Christopher Mathews

## Abstract

**Background:** Little is known regarding the degree to which substance and alcohol use effects on HIV viral suppression are mediated through medication adherence. We hypothesized that the total effects of such use are mediated through adherence.

**Methods:** We included patients with HIV (PWH) receiving care at an urban academic HIV clinic between 2014 and 2018. Eligible patients were those prescribed antiretroviral therapy who completed both patient reported outcome (PRO) questionnaires, and had subsequent plasma viral load (pVL) measurements. Measures included assessments of alcohol use (AUDIT-C), drug use (ASSIST), and self-reported adherence. Substances found in bivariate analysis to predict detectable pVL were modeled separately for mediation effects through adherence. We report natural direct (NDE) and indirect effect (NIE), marginal total effect (MTE) and percentage mediated.

**Results:** Among 3125 Patients who met eligibility criteria, percentages of current use by category were: hazardous alcohol 25.8%, cannabis 27.1%, amphetamines 13.1%, inhalants 11.9%, cocaine 5.3%, sedative-hypnotics 4.5%, opioids 2.9%, and hallucinogens 2.3%. Excellent adherence was reported in 58% and 10% had detectable pVL. Except for sedatives use of other ascertained substances was significantly associated with worse adherence. Bivariate predictors of detectable pVL were [OR(95% CI)]: amphetamine use 2.4 (1.8 -3.2) and opioid use 2.3 (1.3 - 4.0). The percentage mediated by adherence was 36% for amphetamine use, 26.5% for opioid use, and 39% for multiple substance use.

**Conclusion:** Use of amphetamines, opioids, and multiple substances predicted detectable pVL. However, less than 40% of effects were mediated by self-reported adherence.

**Summary:** We examined adherence-mediated effects of hazardous alcohol and substance use on HIV viral suppression. Use of amphetamines, opioids, and multiple substance predicted detectable viral load, however, less than 40% of effects were mediated by self-reported antiretroviral adherence.

## Background

The potential effects of active alcohol and substance use on effectiveness of antiretroviral therapy have been a concern since the advent of antiretroviral therapy. A common assumption has been that any putative effects of alcohol or substance use on HIV viral control would be mediated through effects on antiretroviral medication adherence. (1-3) Regimens with higher resistance barriers, simplified dosing, and long half-lives might attenuate the detrimental effects of alcohol and substance use on both adherence-mediated and adherence-independent causal pathways toward HIV viral load suppression

Some research has suggested potential direct effects of substance use (methamphetamine, cocaine, opioids, cannabis) on immune activation pathways and viral replication. (4-14) Alcohol use has also been associated with potentially detrimental effects on viral control that are independent of medication adherence.(15-18)

We were unable to identify any published clinical studies examining both adherence-mediated and direct effects of hazardous alcohol use and substance use on HIV viral suppression. We hypothesized that adherence is a dominating mediator of alcohol and substance use effects on viral suppression.

## Methods

We conducted a retrospective cross-sectional analysis of persons with HIV (PWH) receiving care in an urban academic HIV clinic between 2014 and 2018. Our cohort included PWH ≥18 years of age on antiretroviral therapy who both completed patient-reported outcomes (PRO) questionnaires and had available the most recent, subsequent HIV plasma viral load (pVL) collected as part of routine care. All participants signed a written consent before study enrollment. The study protocol was approved by UCSD human subjects research protection program (UCSD HRRP #171338)

The PRO questionnaire contained self-report items to ascertain alcohol use, drug use and HIV antiretroviral adherence. Substance use measures included the AUDIT-C and ASSIST as measures of hazardous drinking (AUDIT-C ≥4 in males and ≥ 3 in females) and recent (past 3 months) substance use, respectively.(19, 20) Categories of prescription and recreational drugs surveyed in the PRO battery included: cannabis, amphetamines (including methamphetamine), opioids (both legal and illegal), sedatives-hypnotics, cocaine, inhalants, and hallucinogens. Self-report of adherence was assessed in four ways: (1) the number of doses missed in the last 7 days (1 = 0 missed, 6 = more than 4 missed); (2) Likert scaled adherence (1-6) where 1 = very poor and 6 = excellent; (3) visual analogue scale (VAS) of percent adherence over the past month; (4) last missed dose (1 = within past week, 6 = never skip medications). (1, 21-23)

To select a single measure of medication adherence that most discriminated between those with detectable and undetectable follow-up HIV viral load measurements, we conducted receiver operating characteristic (ROC) analysis of the four candidate measures using binary coded viral load (≤200 copies/ml vs. >200 copies/ml) as reference criterion. To evaluate whether the four adherence measures could jointly enhance discrimination beyond any individual measure, a logistic regression model was fit and predicted probabilities of viral non-suppression were estimated. ROC areas and Hosmer-Lemeshow tests evaluated model discrimination and calibration, respectively. The optimal cut-point of the most discriminating predictor of viral non-suppression was estimated using Stata cutpoint.ado. (StataCorp. 2016. Stata Statistical Software: Release 15.1. College Station, TX).

The selected most discriminating adherence measure was then dichotomized at the optimal cut-point and used in subsequent analyses. Next, we examined bivariate associations: (1) between alcohol and specific substance use and the most discriminating binary measure of adherence; and (2) between alcohol and specific substance use and HIV viral non-suppression. We then examined effects of alcohol and specific substance use on viral suppression in unadjusted and mutually adjusted logistic regression models. Those substances (including alcohol) that were independent predictors of viral non-suppression in logistic models were subsequently selected as independent predictors of viral non-suppression in mediation analyses. Separate mediation models were fit for each selected substance using the Stata paramed.ado program to estimate natural direct effect (NDE), natural indirect effect (NIE) and marginal total effect (MTE) on the odds ratio scale. Percent mediation was estimated as the ratio of natural logarithms of NDE.(24-26) We determined based on the observed prevalence of unsuppressed pVL (10%), prevalence of excellent adherence (58%), and correlation between the drug or alcohol exposure measures and adherence varying from 0.1-0.5, that with type I error = 0.05 and power = 0.8, we could detect a mediation odds ratio from 1.4-1.49, respectively, with a sample size of 3000. (27)

## Results

Table 1 presents study participant characteristics. During the study period, 3125 patients met eligibility criteria for inclusion. The final sample had a median age of 51 years; 54% were non-white, 12% were female, and 72% were men who have sex with men (MSM) as self-reported HIV risk factor. Most (90%) had an undetectable (<200 copies/ml) HIV pVL. The majority of the participants (58%) reported excellent adherence to antiretroviral therapy. The overall prevalence of hazardous alcohol use was 26%. Use of prescription and recreational substances was common. The most used substances were marijuana (27%), amphetamine (13%), inhalants (12%), sedatives-hypnotics (4.5%) and opioids (3%). Multiple substance use was reported in 17% of participant.

**Table 1:** Patient Characteristics.

|  |  |
| --- | --- |
| Sex |  |
| Male | 2742 (87.7%) |
| Female | 383 (12.3%) |
| Race |  |
| White | 1450 (46.4%) |
| Latino | 1025 (32.8%) |
| Black | 417 (13.3%) |
| Asian | 121 (3.9%) |
| Other | 112 (3.6%) |
| HIV Risk Factor |  |
| MSM, not IDU | 2036 (65.2%) |
| IDU, not MSM | 132 (4.2%) |
| IDU+MSM | 208 (6.7%) |
| Heterosexual, not-IDU | 536 (17.2%) |
| All others | 213 (6.8%) |
| Age | median=51.0 (IQR 41.0-58.0) |
| Age | mean=50.1 (sd 11.7) |
| Year |  |
| 2014 | 378 (12.1%) |
| 2015 | 427 (13.7%) |
| 2016 | 574 (18.4%) |
| 2017 | 738 (23.6%) |
| 2018 | 1008 (32.3%) |
| Amphetamines |  |
| No | 2620 (86.9%) |
| Yes | 395 (13.1%) |
| MISSING | 110 observations |
| Sedatives |  |
| No | 2813 (95.5%) |
| Yes | 133 (4.5%) |
| MISSING | 179 observations |
| Cocaine |  |
| No | 2810 (94.7%) |
| Yes | 158 (5.3%) |
| MISSING | 157 observations |
| Opioids |  |
| No | 2884 (97.1%) |
| Yes | 86 (2.9%) |
| MISSING | 155 observations |
| Marijuana |  |
| No | 2177 (72.9%) |
| Yes | 810 (27.1%) |
| MISSING | 138 observations |
| Hallucinogens |  |
| No | 2867 (97.7%) |
| Yes | 66 (2.3%) |
| MISSING | 192 observations |
| Inhalants |  |
| No | 2594 (88.1%) |
| Yes | 349 (11.9%) |
| MISSING | 182 observations |
| AUDIT-C Score | median=1.0 (IQR 0.0-3.0) |
| MISSING | 107 observations |
| AUDIT-C: Hazardous Drinking |  |
| Not Hazardous Drinking | 2240 (74.2%) |
| Hazardous Drinking | 778 (25.8%) |
| MISSING | 107 observations |
| Self Rated Adherence |  |
| < Excellent | 1314 (42.3%) |
| Excellent | 1796 (57.7%) |
| MISSING | 15 observations |
| Self Rated Adherence (VAS) |  |
| <=98 | 927 (55.7%) |
| >98 | 736 (44.3%) |
| MISSING | 1462 observations |
| Interval of last missed dose |  |
| 2-4 weeks, 1-2 weeks, or past week | 678 (39.5%) |
| 1-3 mo, >3 mo, or never | 1040 (60.5%) |
| MISSING | 1407 observations |
| # Missed Doses (7 days) |  |
| ≤ 1 missed dose | 999 (69.7%) |
| > 1 missed dose | 434 (30.3%) |
| MISSING | 1692 observations |
| Detectable pVL (≥200 c/ml) |  |
| Undetectable (<200 c/ml) | 2820 (90.2%) |
| Detectable (≥200 c/ml) | 305 (9.8%) |
| pVL (continuous) median (IQR) | 40 (20,40) |
| Log10 pVL, median (IQR) | 1.6 (1.30, 1.60) |
| Delta(pVLdate - PROdate) | median=24.0 (IQR 2.0-120.0) |
| MISSING | 12 observations |
| Delta(pVLdate - PROdate) | mean=88.8 (sd 154.8) |
| MISSING | 12 observations |
| No Substances Used (ASSIST+AUDIT) |  |
| 0 | 1610 (59.8%) |
| 1 | 633 (23.5%) |
| 2 | 281 (10.4%) |
| 3 | 103 (3.8%) |
| 4-7 | 67 (2.5%) |
| MISSING | 431 observations |
| No Substances Used (ASSIST+AUDIT) | Median = 0.0 (IQR 0.0-1.0) |
| MISSING | 431 observations |
| No Substances Used (ASSIST+AUDIT) | mean=0.7 (sd 1.0) |
| MISSING | 431 observations |

Table 2 presents measurement properties of the four single item adherence measures and the composite predicted adherence proportion based on logistic regression of the four single items on HIV pVL. Because the fewest non-missing values (n=3110) were with item 2 and because the pVL discrimination (ROC area) of this item was highest among the single items adherence measures, we chose this item for use in mediation analysis. The optimal cut-point in predicting pVL was at excellent (6) vs not excellent (≤5).

**Table 2:** PRO Measures of Antiretroviral Medication Adherence: Measurement Properties.

| Item | Content | Possible Score Range | Observed Score Range | Observed Score Median[Mean] | N (observed) | N(missing) | ROC Area<sub>2</sub> | Optimal Cutpoint<sub>2</sub> |
| --- | --- | --- | --- | --- | --- | --- | --- | --- |
| 1. Q303 | # of doses missed (7 days) | 1=0 missed;<br>6=>4 missed | 1--6 | 1 [1.6] | 1433 | 1692 | 0.65 | >1 |
| 2. Q248 | Self Rated Adherence | 1=very poor;<br>6=excellent | 1--6 | 6 [5.2] | 3110 | 15 | 0.71 | >5 |
| 3. Q273 | Self Rated Adherence Visual Analogue Scale (VAS) | 0--100 | 0--100 | 98 [92.3] | 1663 | 1,462 | 0.70 | >98 |
| 4. Q185 | Last Missed Dose | 1=past week;<br>6=never miss | 1--6 | 4 [3.9] | 1718 | 1407 | 0.62 | >3 |
| 5. Composite<sub>1</sub> | Composite logistic regression predictor of viral non-suppression | 0 - 1.0 | 0.05 -- 0.77 | 0.06 [0.10] | 1374 | 1751 | 0.73 | >0.078 |

1. Composite predicted adherence proportion based on logistic regression of items 1-4 on viral load indicator.
2. For items 2-4, undetectable viral load (<200 copies/ml) was coded 1. For items 1 and 5, undetectable viral load was coded 0.

Table 3 presents substance and pVL distributions by self-reported adherence category. Non-adherence was significantly associated with having detectable pVL and with use of amphetamines, cocaine, opioids, marijuana, hallucinogens and inhalants. The Table 4 shows substance use and self-reported adherence by pVL category. Detectable pVL was significantly associated with non-adherence and use of amphetamines and opioids. Hazardous alcohol consumption and use of marijuana, cocaine, inhalants, sedatives-hypnotics, and hallucinogens were not predictive of detectable pVL.

**Table 3:** Distribution of Substance Use by Adherence Level.

| Factor | Level | No Excellent adherence | Excellent adherence | p-value | Test |
| --- | --- | --- | --- | --- | --- |
| N |  | 1323 | 1802 |  |  |
| Amphetamines | No | 1006 (79.5%) | 1614 (92.3%) | <0.001 | Fisher's exact |
|  | Yes | 260 (20.5%) | 135 (7.7%) |  |  |
| Sedatives | No | 1163 (95.0%) | 1650 (95.8%) | 0.32 | Fisher's |
|  |  |  |  |  | exact |
|  | Yes | 61 (5.0%) | 72 (4.2%) |  |  |
| Cocaine | No | 1150 (93.0%) | 1660 (95.9%) | <0.001 | Fisher's exact |
|  | Yes | 87 (7.0%) | 71 (4.1%) |  |  |
| Opioids | No | 1186 (95.9%) | 1698 (98.0%) | 0.001 | Fisher's exact |
|  | Yes | 51 (4.1%) | 35 (2.0%) |  |  |
| Marijuana | No | 848 (68.1%) | 1329 (76.3%) | <0.001 | Fisher's exact |
|  | Yes | 398 (31.9%) | 412 (23.7%) |  |  |
| Hallucinogens | No | 1180 (96.8%) | 1687 (98.4%) | 0.005 | Fisher's exact |
|  | Yes | 39 (3.2%) | 27 (1.6%) |  |  |
| Inhalants | No | 1054 (86.5%) | 1540 (89.3%) | 0.021 | Fisher's exact |
|  | Yes | 165 (13.5%) | 184 (10.7%) |  |  |
| AUDIT-C Score, median (IQR) |  | 1 (0, 4) | 1 (0, 3) | 0.001 | Wilcoxon rank-sum |
| AUDIT-C: Hazardous Drinking | Not Hazardous Drinking | 902 (71.2%) | 1338 (76.4%) | 0.002 | Fisher's exact |
|  | Hazardous Drinking | 364 (28.8%) | 414 (23.6%) |  |  |
| Detectable pVL (≥200 c/ml) | Undetectable (<200 c/ml) | 1101 (83.2%) | 1719 (95.4%) | <0.001 | Fisher's exact |
|  | Detectable (≥200 c/ml) | 222 (16.8%) | 83 (4.6%) |  |  |
| No Substances Used (ASSIST+AUDIT) | 0 | 590 (53.2%) | 1020 (64.3%) | <0.001 | Fisher's exact |
|  | 1 | 281 (25.4%) | 352 (22.2%) |  |  |
|  | 2 | 136 (12.3%) | 145 (9.1%) |  |  |
|  | 3 | 64 (5.8%) | 39 (2.5%) |  |  |
|  | 4-7 | 37 (3.3%) | 30 (1.9%) |  |  |
| No Substances Used (ASSIST+AUDIT), mean (SD) |  | .81 (1.07) | .55 (.90) | <0.001 | Two sample t test |
| No Substances Used (ASSIST+AUDIT), median (IQR) |  | 0 (0, 1) | 0 (0, 1) | <0.001 | Wilcoxon rank-sum |

**Table 4:** Distribution of Substance Use by HIV Viral Suppression Category.

| Factor | Level | Undetectable (<200 c/ml) | Detectable (≥200 c/ml) | p-value | Test |
| --- | --- | --- | --- | --- | --- |
| N |  | 2 820 | 305 |  |  |
| Amphetamines | No | 2 403 (88.1%) | 217(75.6%) | <0.001 | Fisher's exact |
|  | Yes | 3 25 (11.9%) | 70(24.4%) |  |  |
| Sedatives | No | 2 552 (95.5%) | 261(95.3%) | 0.88 | Fisher's exact |
|  | Yes | 1 20 (4.5%) | 13(4.7%) |  |  |
| Cocaine | No | 2 546 (94.7%) | 264(94.3%) | 0.78 | Fisher's exact |
|  | Yes | 142 (5.3%) | 16(5.7%) |  |  |
| Opioids | No | 2 622 (97.4%) | 262(94.2%) | 0.007 | Fisher's exact |
|  | Yes | 70 (2.6%) | 16(5.8%) |  |  |
| Marijuana | No | 1 983 (73.3%) | 194(69.0%) | 0.14 | Fisher's exact |
|  | Yes | 723 (26.7%) | 87(31.0%) |  |  |
| Hallucinogens | No | 2 607 (97.9%) | 260(96.3%) | 0.13 | Fisher's exact |
|  | Yes | 56 (2.1%) | 10(3.7%) |  |  |
| Inhalants | No | 2 360 (88.3%) | 234(87.0%) | 0.55 | Fisher's exact |
|  | Yes | 314 (11.7%) | 35(13.0%) |  |  |
| AUDIT-C Score, median (IQR) |  | 1 (0, 3) | 1(0,3) | 0.64 | Wilcoxon rank-sum |
| AUDIT-C: Hazardous Drinking | Not Hazardous Drinking<sup>1</sup> | 2 020 (74.2%) | 220(74.3%) | 1.00 | Fisher's exact |
|  | Hazardous Drinking<sup>2</sup> | 702 (25.8%) | 76(25.7%) |  |  |
| ART Adherence | No Excellent adherence<sup>3</sup> | 1 101 (39.0%) | 222(72.8%) | <0.001 | Fisher's exact |
|  | Excellent adherence<sup>4</sup> | 1 719 (61.0%) | 83(27.2%) |  |  |
| No Substances Used (ASSIST+AUDIT), mean (SD)<sup>5</sup> |  | .64 (.97) | .86(1.09) | <0.001 | Two sample t test |
| No Substances Used (ASSIST+AUDIT), median (IQR)<sup>6</sup> |  | 0 (0, 1) | 0(0,1) | <0.001 | Wilcoxon rank-sum |

1. AUDIT-C<3 in female and <4 men 2. AUDIT-C ≥3 in female and ≥4 in men 3. Adherence self-report either: poor, fair, good, very good 4. Adherence self-report: excellent 5. SD: standard deviation 6. IQR: interquartile range

In both unadjusted and mutually adjusted logistic regression models, use of amphetamines and use of opioids were significantly associated with having a detectable HIV viral load (Table 5). Those reporting amphetamine or opioid use were more than twice as likely to have detectable pVL. The failure of binary coded hazardous alcohol use to predict virology non-suppression was confirmed in ROC analysis of the numeric AUDIT-C score evaluated against pVL as reference criterion (ROC 0.492, 95% C.I. 0.457 - 0.527).

**Table 5:** Unadjusted and Adjusted Effects of Drug and Alcohol Use on Detectable HIV Viral Load.

| Predictor | Unadjusted Effects |  |  | Mutual Adjusted Effects<sub>1</sub> |  |  |
| --- | --- | --- | --- | --- | --- | --- |
|  | Odds Ratio | 95% CI<sub>2</sub> | p-value | Odds Ratio | 95% CI | p-value |
| Sedatives | 1.059 | 0.589, 1.904 | 0.847 | 0.693 | 0.352, 1.366 | 0.29 |
| Cocaine | 1.087 | 0.638, 1.850 | 0.76 | 0.568 | 0.287, 1.124 | 0.104 |
| Amphetamines | 2.385 | 1.779, 3.198 | 0.0001 | 2.166 | 1.509, 3.107 | <0.0001 |
| Opioids | 2.287 | 1.310, 3.996 | 0.004 | 2.173 | 1.157, 4.081 | 0.016 |
| Marijuana | 1.23 | 0.942, 1.606 | 0.128 | 1.028 | 0.757, 1.396 | 0.862 |
| Hallucinogens | 1.791 | 0.903, 3.551 | 0.095 | 1.46 | 0.65, 3.281 | 0.359 |
| Inhalant | 1.124 | 0.773, 1.634 | 0.54 | 0.807 | 0.521, 1.25 | 0.337 |
| Alcohol:Hazardous | 0.994 | 0.755, 1.308 | 0.966 | 1.044 | 0.767, 1.421 | 0.783 |

1. Hosmer-Lemeshow chi-square (7) = 4.03, p = 0.545. ROC area = 0.591
2. CI: confidence interval

Table 6 presents mediation model results for those substances found to be significant predictors of pVL non-suppression. For single substances, the percent mediation was highest (36.2%) for amphetamine use, while for opioids, the percent mediated was 26.5%. The percentage mediation for multiple substance use was 39.7%. Patients with no missing values (complete cases) for mediation analyses (n = 2921) did not differ from incomplete (excluded) patients by year of measurement, sex, or age (p>0.10). However, patients with non-missing values were more likely to identify as white verse non-white (47% vs. 37%, p = 0.002) and to report male sex with men, and not injection drug use (MSM, not-IDU) as their HIV transmission risk factor (66% vs. 57%, p = 0.028).

**Table 6:** Mediation Analysis Results.

|  |  | Effect Odds |  |  |  |
| --- | --- | --- | --- | --- | --- |
|  |  | Ratio | Bootstrap Std Error | 95% C.I. |  |
| Amphetamines |  |  |  |  |  |
|  | NDE | 1.75 | 0.28 | 1.26 | 2.38 |
|  | NIE | 1.37 | 0.05 | 1.29 | 1.48 |
|  | MTE | 2.40 | 0.38 | 1.75 | 3.31 |
|  | Percent Mediated | 36.18% |  |  |  |
| Opioids |  |  |  |  |  |
|  | NDE | 1.86 | 0.60 | 0.86 | 2.99 |
|  | NIE | 1.25 | 0.08 | 1.11 | 1.41 |
|  | MTE | 2.33 | 0.75 | 1.10 | 3.83 |
|  | Percent Mediated | 26.48% |  |  |  |
| No. Substances Reported |  |  |  |  |  |
|  | NDE | 1.13 | 0.07 | 0.98 | 1.28 |
|  | NIE | 1.09 | 0.02 | 1.06 | 1.12 |
|  | MTE | 1.23 | 0.08 | 1.07 | 1.39 |
|  | Percent Mediated | 39.74% |  |  |  |

NDE: natural direct effect, NIE: natural indirect effect, MTE: marginal total effect

## Discussion

Our research found that recent use of amphetamines, cocaine, opioids, marijuana, hallucinogens, inhalants, hazardous alcohol use (AUDIT-C), and multiple substances use were significantly associated with less than excellent self-reported antiretroviral adherence. Although not entirely uniform in their conclusions, many previous studies have documented detrimental effects of use of specific recreational substances and alcohol on various measures of adherence that included self-report or electronic monitoring approaches. (2, 3, 28-32) Most robust conclusions pertain to the use of amphetamines, cocaine, opioids, and alcohol. Regarding cannabis use, intensity and regularity of use may modify effects on adherence. (33, 34) Our focus, however, was on distal and more impactful outcomes of antiretroviral therapy, specifically HIV viral suppression. We were surprised to find that only self-reported use of amphetamines and of opioids as individual substances were predictive of viral non-suppression while use of multiple substances was also predictive. Differences in study design and instruments used to measure hazardous alcohol use could account for differences with other studies that observed that hazardous alcohol use is associated with having detectable viral loads. Most of these studies used cohort designs. (2, 35, 36) However, a prospective randomized control trial failed to show the same association. (37) In many of these cohorts, either exposure was defined as any alcohol use or other instruments than AUDIT-C were used. (38) Yet, our findings mirror findings of a previous study which found that hazardous alcohol use was associated with ART non-adherence but not with HIV pVL. (39) We believe that our failure to detect an effect for hazardous alcohol use was not attributable to the specific AUDIT-C cutoff score used because the ROC analysis using full range numeric AUDIT-C score showed that the measure did not discriminate between those with and without viral suppression.

Mediation analyses restricted to those substances that were associated with plasma viral load non-suppression (amphetamines, opioids, and multiple substances use) yielded a result that was not anticipated. Less than 40% of the effects on viral non-suppression appeared to be mediated through our measure of medication adherence. There are several potential explanations for this finding. It could represent a true finding pointing to the effect of these substances not mediated through antiretroviral adherence. Another potential explanation is the occurrence of pharmacokinetic or pharmacodynamics drug-drug interactions that adversely influenced antiretroviral effects independently of adherence behaviors. (40) Other possible explanations have to do with measurement issues: the specific metric chosen for substance use (any past 3 month use on NIDA ASSIST); the timing of substance use recall period with respect to subsequent viral load measurement (median 24 days); use of an insensitive measure of medication adherence (although we selected the most discriminating of the candidate measures available [Table 2]). Residual confounding and collider effects resulting in selection bias due to missing values must also be considered. These issues merit further exploration and replication efforts.

Additional limitations of our findings are that they represent a single-center experience, a cross-sectional rather than longitudinal design, and are based on a data set with missing data that limited complete case analysis for multivariable components of the analysis. Our analysis included mainly whites and MSM and our results might not generalize to HIV with diverse characteristics. Nonetheless, we conclude that the mediation analysis findings are novel and provocative, meriting examination in expanded cohorts with longitudinal follow-up.

## Data Availability

All data produced in the present study are available upon reasonable request to the authors

